# Twitter Engagement of U.S. Psychiatry Residency Programs with Black Lives Matter and Coronavirus Disease 2019 (COVID-19)

**DOI:** 10.1101/2020.10.20.20215723

**Authors:** Osama El-Gabalawy, Candice J. Kim, Amanda V. Chen, Shaan Kamal

## Abstract

Social media have become popular platforms to disseminate information, especially related to politicized topics such as BLM and COVID-19. To better understand how medical institutions have engaged with the social media discourse on BLM and COVID-19, we examined psychiatry residency programs’ tweets in response to George Floyd’s murder and during the first 6 months of the COVID-19 pandemic in the U.S. Only 14% of the 249 evaluated psychiatry residency programs had Twitter accounts (we included programs with their own account or their affiliated psychiatry department account) indicating a substantial absence on social media. Of those programs, 78% tweeted at least once about COVID-19 (1,153 tweets) and 56% tweeted at least once about the BLM movement (117 tweets). The top three purposes of tweets were sharing media, posting about an event, and sharing a resource.

## Introduction

Concerns about mental health have been raised across the nation in the wake of the Black Lives Matter (BLM) movement and the coronavirus disease 2019 (COVID-19) pandemic.1-4 Social media, particularly Twitter, have become popular platforms to disseminate information, especially related to politicized topics such as BLM and COVID-19. To better understand how medical institutions have engaged with the social media discourse on BLM and COVID-19, we examined psychiatry residency programs’ tweets in response to George Floyd’s murder and during the first 6 months of the COVID-19 pandemic in the U.S.

## Methods

A list of 249 US psychiatry residency programs was taken from Doximity.com in November, 2019. We determined social media presence for each program by searching each program’s website and using a Google search to identify Twitter accounts of the residency program or its affiliated psychiatry department. All tweets and re-tweets from January 1st to June 29, 2020 were scraped from the identified Twitter accounts. We established BLM & COVID-19 search terms (see supplement section for lists) by starting with a minimal set (e.g. “black lives matter” for BLM and “COVID” for COVID-19), and iterating to include commonly associated search terms found in the positive tweets (e.g. “quarantine”). We used COVID-19 search terms on tweets from January 1 to June 29, 2020 and BLM search terms on tweets from May 25 (date of George Floyd’s murder) to June 29, 2020. Of the subset of tweets positive for a COVID-19 search term, we conducted a subsequent search using BLM terms to identify intersectional tweets. Two of us (C.J.K. and A.V.C.) reviewed the tweets and independently coded the data, generating a list of themes using a grounded theory approach. The two coders iteratively compared and refined themes to produce a final set of themes. Word and Excel (Microsoft Corp) was used to manage the data.

## Results

36 out of the 249 psychiatry residency programs in the U.S. had a Twitter account. Of these programs, only 55.6 % (20/36) tweeted at least once about the BLM movement, while 77.8% (28/36) tweeted at least once about the COVID-19 pandemic during the study period for each subset of tweets (May 25 to June 29, 2020 for BLM; January 1 to June 29, 2020 for COVID-19). A total of 117 tweets were positive for BLM key words. 2 of these tweets were removed because their content was not directly related to BLM. 1,153 tweets were positive for COVID-19 key words; only 16 of these tweets were related to BLM. For our subsequent qualitative analyses, we included all BLM-related tweets and a 10% random sample of the COVID-19 related tweets for a total of 230 tweets, with 115 tweets for BLM and COVID-19 each.

Overall, the top three purposes of tweets were sharing media (43.5%, 100/250), posting about an event (24.8%, 57/250), and sharing a resource (13.9%, 32/230) (Table 1). Of the tweets that shared media, the majority of them shared news articles (75%, 75/100) rather than videos (13.0%, 13/100), podcast or radio episodes (6.0%, 6/100), or journal articles (6.0%, 6/100) (Table 2). Of the tweets that posted about an event, the majority posted about a protest or vigil (43.9%, 25/57) or a talk (31.6%, 18/57) (Table 3). Notably, only two tweets that posted about an event were publicizing a training session (3.5%, 2/57) (Table 3).

Comparing the BLM and COVID-19 tweets, several key differences emerged. The COVID-19 subset had close to 7 times as many tweets than the BLM subset sharing a resource (24.4%, 28/115, vs. 3.5%, 4/115) (Table 1). However, the BLM subset had over 5 times as many tweets than the COVID-19 subset posting about an event (41.7%, 48/115, vs. 7.8%, 9/115) (Table 1). More than half of the BLM tweets posting about an event were publicizing a protest or vigil (52.1%, 25/48).

## Discussion

Few psychiatry residency programs have an online presence through Twitter. Of those that do, more programs tweeted at least once about COVID-19 than BLM. Tweets primarily focused on sharing media, posting about an event, or sharing a resource. However, very few COVID-19 tweets had BLM-related content, and even fewer yet discussed the pandemic’s disproportionate impact on Black communities and their mental health. Limitations include taking a 10% sample of COVID-19 tweets for qualitative analysis rather than a complete sample; and including BLM tweets for the month after George Floyd’s murder although the BLM movement has continued past the end of June. Nevertheless, this study provides insight on how medical institutions, specifically residency programs, engage with current political discourse through social media. Findings from our study highlight the need to share resources to support mental health of Black communities through accessible forms of communication like Twitter. Future research is needed to better understand long-term engagement on social media by medical institutions regarding public health crises like police brutality and the pandemic.

## Data Availability

All data in the manuscript is publicly available from the sources described, our work only provides an analysis for it. 

https://www.twitter.com

## Author Contributions

OE and SK contributed to the compilation of all data. OE, CK, and AC performed the analysis of the data and writing of the manuscript. All authors edited and approved of the final manuscript.

## Figures

**Table. Primary Purposes for Programs' Tweets Related to Black Lives Matter or COVID-19**
| Primary Purpose | No (%) |  |  |
| --- | --- | --- | --- |
|  | Black Lives Matter<br>(n = 115) | COVID-19<br>(n = 115) | Overall<br>(n = 230) |
| Sharing media | 37 (32.2) | 63 (54.8) | 100 (43.5) |
| News article | 28 (24.3) | 47 (40.9) | 75 (32.6) |
| Video | 3 (2.6) | 10 (8.7) | 13 (5.7) |
| Podcast/radio | 3 (2.6) | 3 (2.6) | 6 (2.6) |
| Journal/research article | 3 (2.6) | 3 (2.6) | 6 (2.6) |
| Posting about event | 48 (41.7) | 9 (7.8) | 57 (24.8) |
| Protest/vigil | 25 (2.2) | 0 (0.0) | 25 (10.9) |
| Talk | 10 (8.7) | 8 (7.0) | 18 (7.8) |
| Town hall | 7 (6.1) | 0 (0.0) | 7 (3.0) |
| Juneteenth | 5 (4.3) | 0 (0.0) | 5 (2.2) |
| Training | 1 (0.9) | 1 (0.9) | 2 (0.9) |
| Sharing resource | 4 (3.5) | 28 (24.4) | 32 (13.9) |
| Mission statement | 17 (14.8) | 0 (0) | 17 (7.4) |
| Celebrating people | 3 (2.6) | 6 (5.2) | 9 (3.9) |
| Sharing clinic information | 0 (0) | 7 (6.1) | 7 (3.0) |
| Sharing facts | 4 (3.5) | 1 (0.9) | 5 (2.2) |
| Call to action | 2 (1.7) | 1 (0.9) | 3 (1.3) |

## Supplemental Section

### BLM Search terms

1. black lives matter
2. black lives
3. black
4. blacklivesmatter
5. blacklives
6. blm
7. African American
8. African-American
9. ahmaud arbery
10. breonna taylor
11. george floyd
12. breonnataylor
13. georgefloyd
14. amaudarbery,
15. injustice
16. racism
17. police brutality
18. disparity
19. disparities

### COVID Search Terms

1. covid
2. coronavirus
3. social distancing
4. pandemic
5. quarantine

## References

1. Pfefferbaum B, North CS. Mental Health and the Covid-19 Pandemic. N Engl J Med. April 2020. doi:10.1056/nejmp2008017

2. Reger MA, Stanley IH, Joiner TE. Suicide Mortality and Coronavirus Disease 2019—A Perfect Storm? JAMA Psychiatry. April 2020. doi:10.1001/jamapsychiatry.2020.1060

3. Bor J, Venkataramani AS, Williams DR, Tsai AC. Police killings and their spillover effects on the mental health of black Americans: a population-based, quasi-experimental study. Lancet. 2018;392(10144):302–310. doi:10.1016/S0140-6736(18)31130-9

4. Egede LE, Walker RJ. Structural Racism, Social Risk Factors, and Covid-19 — A Dangerous Convergence for Black Americans. N Engl J Med. July 2020. doi:10.1056/nejmp2023616

